# Dynamic Stroke Risk Stratification via Machine Learning: A Multi-Level Single-Center Study

**DOI:** 10.64898/2025.12.03.25341598

**Authors:** Zhirui Liao, Zesheng Yan, Jing Huang, Ximei Chen, Yinghua Yang

**Affiliations:** School of Public Health, Wuhan University, Wuhan 430072, Hubei Province, P.R. China; Center for Disease Control and Prevention of Qingshanhu District, Nanchang City, Nanchang 330038, Jiangxi Province, P.R. China

**Keywords:** Stroke, Risk Prediction, Machine Learning, Dynamic Features, Longitudinal Cohort

## Abstract

**Background:** Stroke is a leading global public health challenge and the second leading cause of death worldwide. In China, its burden continues to escalate amid population aging and a growing prevalence of unhealthy lifestyles. Traditional static stroke risk prediction models, constrained by cross-sectional data, fail to capture dynamic changes in physiological parameters and behavioral factors, resulting in inherent limitations. This study therefore aimed to construct and validate a multi-tiered dynamic stroke risk prediction system.

**Methods:** A single-center longitudinal prospective cohort study (2018–2022) enrolled community-dwelling populations and outpatients who completed three consecutive follow-ups. Three sequential machine learning models were developed, targeting general population screening, high-risk population refinement, and longitudinal population monitoring, respectively. Stratified cross-validation was used for model validation (10-fold for Model 1, 5-fold for Models 2 and 3), with the area under the curve (AUC) as the primary evaluation metric.

**Results:** Model 1 (for general population high-risk conversion prediction) achieved an AUC of 0.835, which significantly reduced the missed detection rate of individuals with “normal static indicators but abnormal dynamic trends.” Model 2 (for high-risk population stroke onset prediction) had LGB_Conservative as its optimal algorithm, with an AUC of 0.8479. Model 3 (for longitudinal population dual-outcome prediction) showed an AUC of 0.761, with stroke event rates of 3.1% in the low-risk group and 95.2% in the very high-risk group.

**Conclusion:** This multi-tiered dynamic prediction system effectively addresses the limitations of traditional static models, yet requires external validation using multi-center data to confirm its generalizability. It provides a novel tool for personalized stroke prevention in clinical and public health practice.

## Introduction

Stroke is the second leading cause of death globally and a leading cause of long-term mortality and disability, exerting an enormous global public health burden**^Error!^ ^Reference^ ^source^ ^not^ ^found^**. In China, population aging and the rising prevalence of unhealthy lifestyles are steadily exacerbating this burden **Error! Reference source not found.** Given stroke’s high disability and mortality rates, coupled with its profound economic consequences, developing robust risk prediction models to enable precise primary prevention has become an urgent priority in both public health and clinical practice.

**Figure 1.**
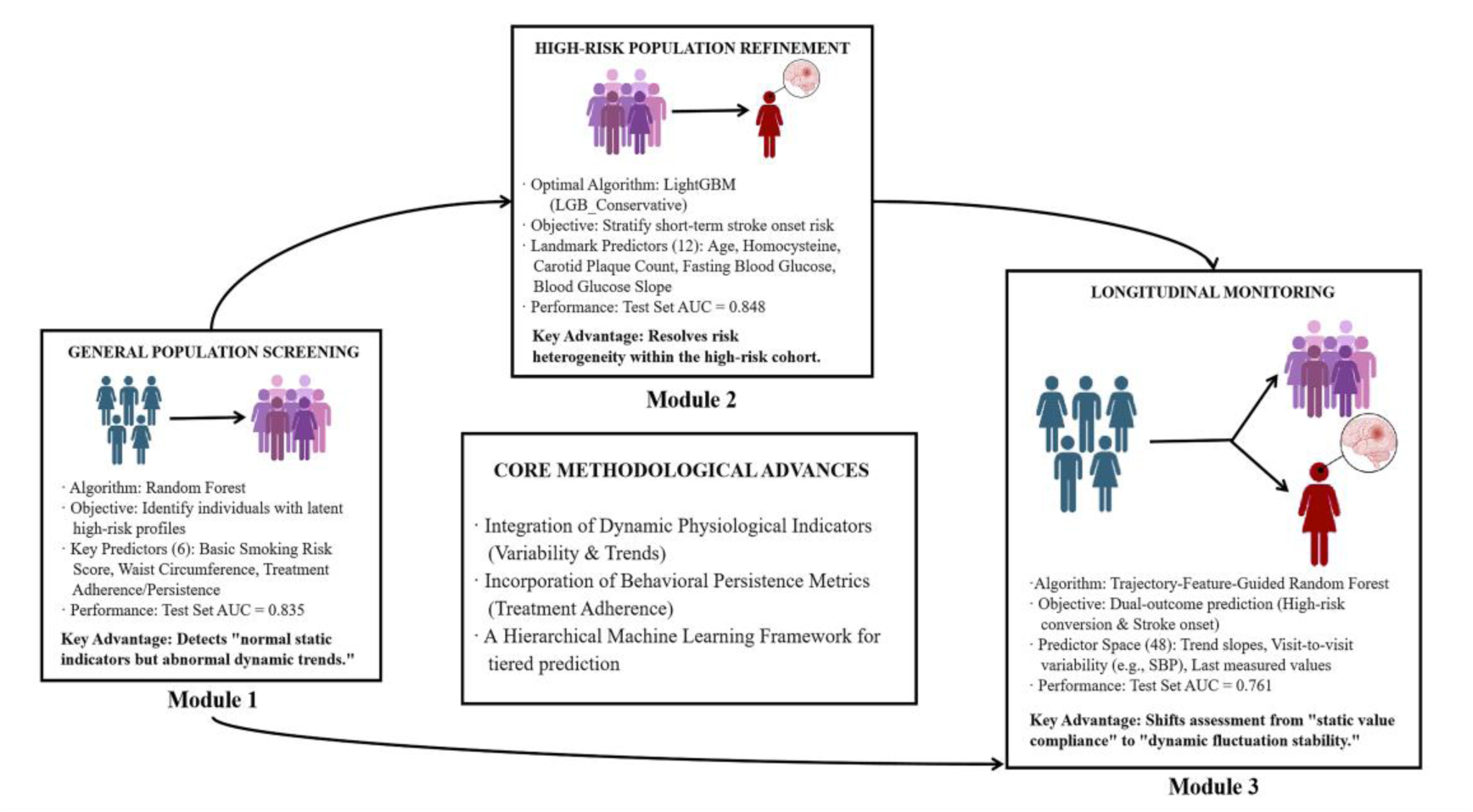
Schematic of the study’s dynamic risk stratification system, developed and validated in a single-center prospective cohort. The framework leverages machine learning models that incorporate dynamic parameters and trajectory features for general screening, high-risk refinement, and longitudinal monitoring.

Current widely used stroke risk screening strategies—both domestically and internationally—primarily rely on cross-sectional assessment of static risk factors (e.g., hypertension, atrial fibrillation, smoking) [2]. While these static models have played a historical role in rapid population-level screening^[3]^, their inherent limitations are increasingly prominent: they provide only a “static snapshot” and fail to capture the temporal dynamic evolution of risk factors. Consequently, they risk overlooking the critical transition from quantitative to qualitative shifts in risk^[4][5]^.

The key to addressing this limitation lies in a more nuanced understanding of dynamic fluctuations in physiological parameters. Cutting-edge cardiovascular research has confirmed that long-term visit-to-visit variability in parameters such as blood pressure and blood glucose is a more robust predictor of cardio-cerebrovascular events than their mean values—and this predictive power is independent of those average levels**^Error!^ ^Reference^ ^source^ ^not^ ^found.^**^[7]^. Furthermore, the persistence of behavioral factors (e.g., long-term treatment adherence, lifestyle stability) collectively forms a more clinically relevant individual risk profile than single-time-point evaluations**^Error!^ ^Reference^ ^source^ ^not^ ^found.^**. Despite compelling evidence supporting their value, most existing stroke prediction models have failed to systematically integrate these dynamic metrics and behavioral persistence indicators into a holistic assessment framework.

Additionally, substantial heterogeneity exists within the already identified high-risk population^[9][10]^. Currently, there is a paucity of effective tools to further precisely identify individuals at imminent high risk of stroke onset within this subgroup—leading to the suboptimal allocation of preventive healthcare resources. Clinical practice now requires not merely an abstract risk probability, but a predictive tool that can detect individual-specific abnormal data fluctuation patterns and offer an early intervention window supported by pathophysiological reasoning.

To fill these gaps, this study seeks to utilize data from a single-center longitudinal prospective cohort to develop and validate a novel multi-tiered dynamic stroke risk prediction system. This system consists of three logically sequential core models:

1. An initial screening model for the general population, which redefines traditional static high-risk identification by integrating dynamic trends and behavioral metrics from multiple follow-up visits;
2. A precision screening model for the identified high-risk subgroup, which integrates both static and dynamic parameters to predict short-term stroke onset risk and enable risk re-stratification;
3. An individualized longitudinal monitoring model, which can simultaneously alert to both high-risk conversion and stroke onset by detecting clinically meaningful abnormal trajectory patterns—ultimately guiding intervention strategies.

We hypothesize that this framework will facilitate earlier risk stratification, more accurate risk prognostication, and ultimately deliver actionable decision support to advance personalized stroke prevention.

## Methods

### 1. Data Source and Preprocessing

A single-center longitudinal prospective cohort design was adopted. Data were derived from community-dwelling populations and outpatients who consecutively completed three follow-up visits between 2018 and 2022. Based on the research objectives, the population was divided into three independent modeling cohorts:

- Model 1 (Prediction of high-risk conversion in the general population): A total of 2486 individuals with complete data from three follow-up visits were included, and they were stratified randomly into a training set and a test set at a ratio of 7:3.
- Model 2 (Prediction of stroke onset in high-risk populations): 592 high-risk stroke individuals with complete carotid ultrasound data were included, and stratified randomly divided into a training set and a test set at a 75:25 ratio.
- Model 3 (Longitudinal trajectory prediction): 2,486 participants with complete dual outcome data (high-risk conversion and stroke onset) were enrolled, and stratified randomly split into a training set and a test set at an 8:2 ratio.

All participants signed informed consent forms, and the study was conducted in accordance with Good Clinical Practice (GCP) guidelines. This study was approved by the Ethics Committee of Xuanwu Hospital, Capital Medical University (Approval No.: Clinical Research Review [2015] No. 024).

### 2. Data Preprocessing and Feature Engineering

#### 2.1 Data Preprocessing

A standardized process was used for data handling:

- Missing numerical values were imputed with the median, while missing categorical values were filled with the mode^[11]^.
- Binary variables were subjected to 0/1 encoding, and multicategorical variables were processed with label encoding or one-hot encoding depending on model requirements.
- To address class imbalance, class weight balancing was applied during the training of Models 1 and 2; additionally, the scale_pos_weight parameter was adjusted specifically for the XGBoost algorithm in Model 2^[12]^.
- A fixed random seed (random_state=42) was set for all steps to ensure reproducibility**^Error!Reference source not found^**.
- For the protection of participants’ personal information, in accordance with the Approval Opinion of the Ethics Committee of Xuanwu Hospital, Capital Medical University (Approval No.: Clinical Research Review [2015] No. 024), all participant data were de-identified prior to data analysis. Specifically, direct identifiers such as name, ID number, contact phone number, and residential address were permanently removed from the dataset; indirect identifiers (e.g., age range, residential community) were pseudonymized to prevent reverse identification. All de-identified data were stored on an encrypted server, with access permissions restricted to core members of the research team; data transmission was protected using the Secure File Transfer Protocol (SFTP) to avoid information leakage.

**Figure 2.**
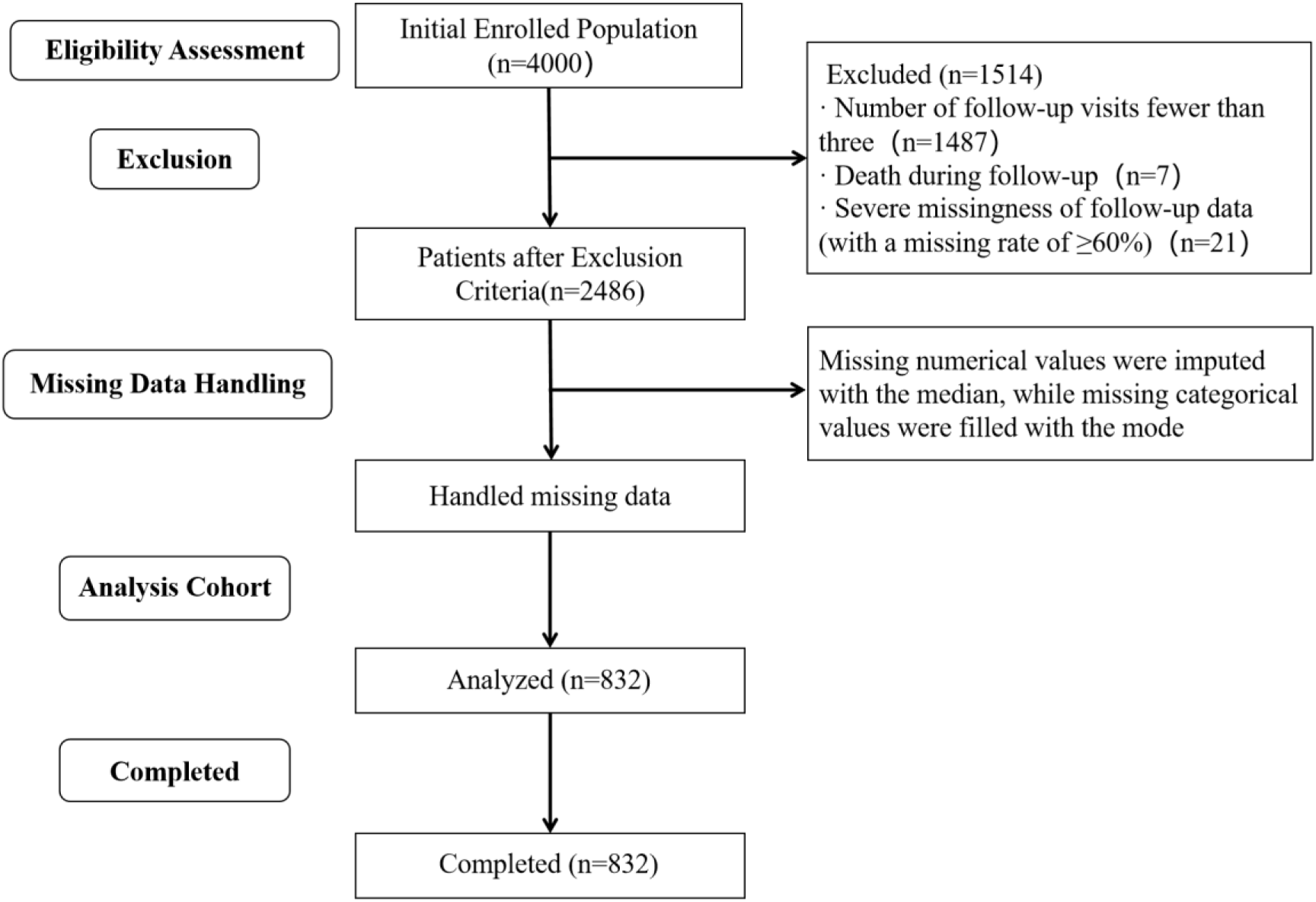
Flowchart of participant inclusion and exclusion for the screening cohort.

#### 2.2 Feature Construction

Three categories of key features were constructed to enhance predictive performance:

- Treatment Persistence Score: Treatment persistence was quantified by integrating medication type, persistence ratio (calculated as [regular medication duration / follow-up duration]×100, excluding unplanned ≥ 7-day drug interruption), and adjustment for temporary adherence fluctuations.
- Treatment adherence: Defined as the proportion of self-reported compliance with prescribed medications at the time of treatment indication. It was incorporated into the models as “Antihypertensive Adherence Grade,” “Hypoglycemic Adherence Grade,” and “Lipid-Lowering Adherence Grade.”[14]
- Basic Smoking Risk Score: Tobacco exposure was quantified by integrating smoking status, pack-years (calculated as [cigarettes per day / 20] × years of smoking), and the risk attenuation effect in former smokers. It was included as a single feature named “Basic Smoking Risk Score.” [15]
- Genetic burden score for family history of diseases: 1 point was assigned for a history of stroke or coronary heart disease (CHD) in either parent; weighted scoring was applied for affected siblings (base score = 0.5, increasing logarithmically with the number of affected siblings). Participants with no relevant family history received a score of 0. Derived features included “CHD

Family History Score,” “Diabetes Family History Score,” and “Hypertension Family History Score.”Error! **^Reference source not found^**.

#### 2.3 Feature Selection Strategy

A conservative feature selection strategy was adopted for all models to maximize simplicity while ensuring predictive performance:

- Model 1: The initial feature set included 27 new variables (variability-processed data and other potential predictors), excluding traditional static model factors. Features with non-zero coefficients were screened using strongly regularized L1-penalized logistic regression[17], and feature importance was calculated via a conservative-parameter Random Forest. Pearson correlation coefficients between features and outcome variables were computed, and only highly correlated features were retained, with the final number of features strictly limited to 6.
- Model 2: The initial feature set was expanded to 92 variables, supplementing static data and carotid ultrasound indicators to the variables of Model 1. Features were filtered using L1-regularized logistic regression, and feature importance was evaluated with a conservative-parameter Random Forest. The top 12 key features were selected based on combined importance ranking.
- Model 3: The initial set included 48 trajectory features, and automatic selection was performed using the built-in importance assessment of the Random Forest algorithm.

### 3. Prediction Model Construction

#### 3.1 Model Framework and Algorithm Selection

Three models were designed with targeted frameworks based on distinct research objectives:

- Model 1: Adopted a “feature selection-single model optimization” framework, using the Random Forest algorithm.
- Model 2: Employed a “multi-algorithm comparison” framework, with parallel training of 7 algorithms: 2 regularized logistic regressions (L1, L2), 2 parametric gradient Random Forests (Conservative, Simple), GBM, XGBoost, and LightGBM. The optimal algorithm was selected for final analysis.
- Model 3: Utilized a “trajectory feature construction-single model focus” framework, using the Random Forest algorithm to simultaneously predict dual outcomes.

#### 3.2 Model Training and Validation

- All models were validated using stratified cross-validation to assess generalizability: 10-fold for Model 1, and 5-fold for Models 2 and 3.
- Hyperparameter optimization was performed via grid search. The Area Under the Curve (AUC) was used as the primary evaluation metric for model performance and selection. For Model 3, the Precision-Recall Area Under the Curve (PR-AUC) was additionally calculated.

#### 3.3 Overfitting Assessment

The degree of overfitting was quantified by comparing the AUC difference between the training and test sets. A tolerance threshold of 0.05 was set for Model 1, while a stricter threshold of 0.02 was applied for Models 2 and 3. This difference was incorporated as a penalty term in the algorithm selection process for Model 2.

All analyses were performed using Python, and the study followed the Transparent Reporting of a multivariable prediction model for Individual Prognosis Or Diagnosis (TRIPOD) reporting guidelines.

## Results

### 1. Baseline Characteristics of the Study Population

The baseline characteristics of participants included in the three models are summarized below. All quantitative data are presented as mean±standard deviation, and categorical data as n (percentage). Intergroup comparisons were performed using the independent samples t-test (for quantitative data) or chi-square (χ²) test (for categorical data), with statistical significance defined as P < 0.05.

The baseline characteristics of the participants across the three modeling cohorts are summarized in Table 1. Critically, the cohort for Model 2 (high-risk stroke population) exhibited a significantly more adverse cardiovascular risk profile compared to the general population in Model 1, characterized by advanced age, elevated systolic blood pressure, higher fasting blood glucose, and homocysteine levels (all P < 0.05). This gradient in baseline risk validates the cohort stratification strategy and underscores the necessity of tiered prediction models.

**Table 1.**
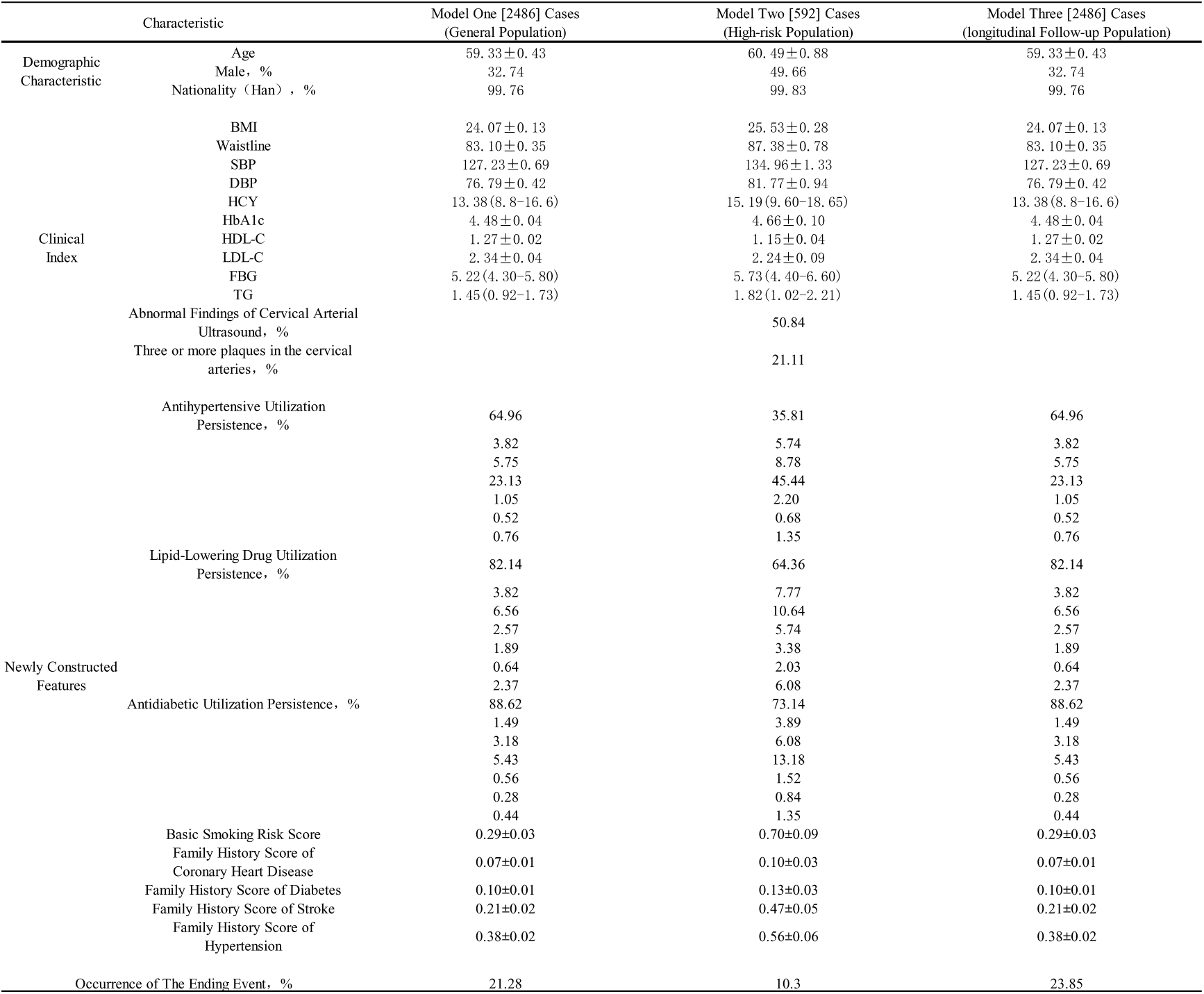
Baseline Characteristics of Participants in the Three Modeling Cohorts. Note: Quantitative variables are presented as mean±standard deviation or median (interquartile range), as appropriate. Categorical variables are expressed as n (%). Abbreviations: BMI, body mass index; SBP, systolic blood pressure; DBP, diastolic blood pressure; HCY, homocysteine; HbA1c, glycated hemoglobin; HDL-C, high-density lipoprotein cholesterol; LDL-C, low-density lipoprotein cholesterol; FBG, fasting blood glucose; TG, triglycerides. Medication persistence levels are ranked ordinally, with higher levels indicating better persistence.

**Table 2.** Risk Stratification and Event Rates in Model 3 According to Predicted Risk Categories. Note: Participants were stratified into four risk groups based on Model 3 predictions. Event rates represent the proportion of individuals experiencing either high-risk conversion or stroke onset within each stratum.

| Risk Level | Sample Size | Event Rate | Mean Risk Score |
| --- | --- | --- | --- |
| Low-Risk | 1746 | 3.1% | 0.134 |
| Moderate-Risk | 493 | 46.0% | 0.311 |
| High-Risk | 123 | 84.6% | 0.431 |
| Very High-Risk | 124 | 95.2% | 0.572 |

### 2. Evaluation of Model Predictive Performance

#### 2.1 Model 1 (Prediction of Stroke High-Risk Conversion in the General Population)

This model ultimately retained 6 core features: Basic Smoking Risk Score, waist circumference, Antihypertensive Treatment Persistence, Hypoglycemic Treatment Persistence, Antihypertensive Adherence Grade, and high-density lipoprotein cholesterol (HDL-C).

- In the test set, the model achieved an Area Under the Curve (AUC) of 0.835, with a precision of 0.50 and a recall (sensitivity) of 0.74. Notably, it significantly reduced the missed detection of individuals with “normal static indicators but abnormal dynamic trends”—a key advantage over traditional static models.
- The AUC difference between the training and test sets was only −0.003, indicating negligible overfitting and strong generalizability.
- Core hyperparameters of the model were set as follows: n_estimators = 30, max_depth = 5, min_samples_split = 30, min_samples_leaf = 10, max_features = 0.3, and class_weight = “balanced”.

**Figure 3.**
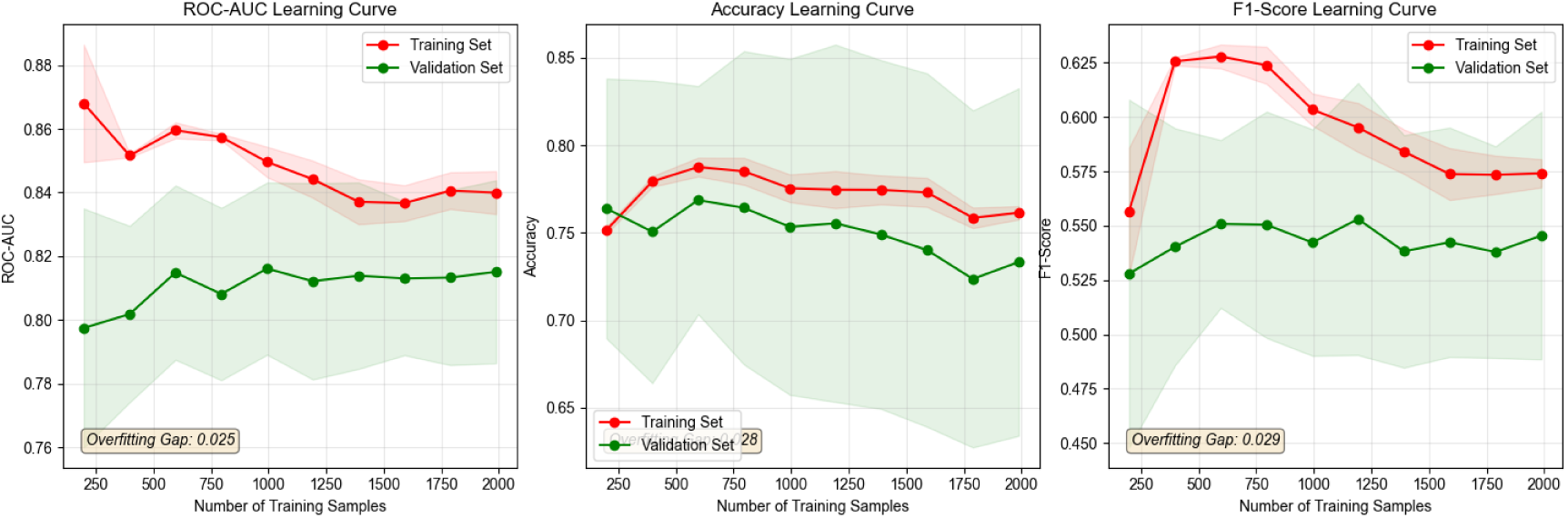
Learning curves for Model 1 (prediction of high-risk conversion in the general population), depicting the area under the curve (AUC), accuracy, and F1-score across training and test sets.

**Figure 4.**
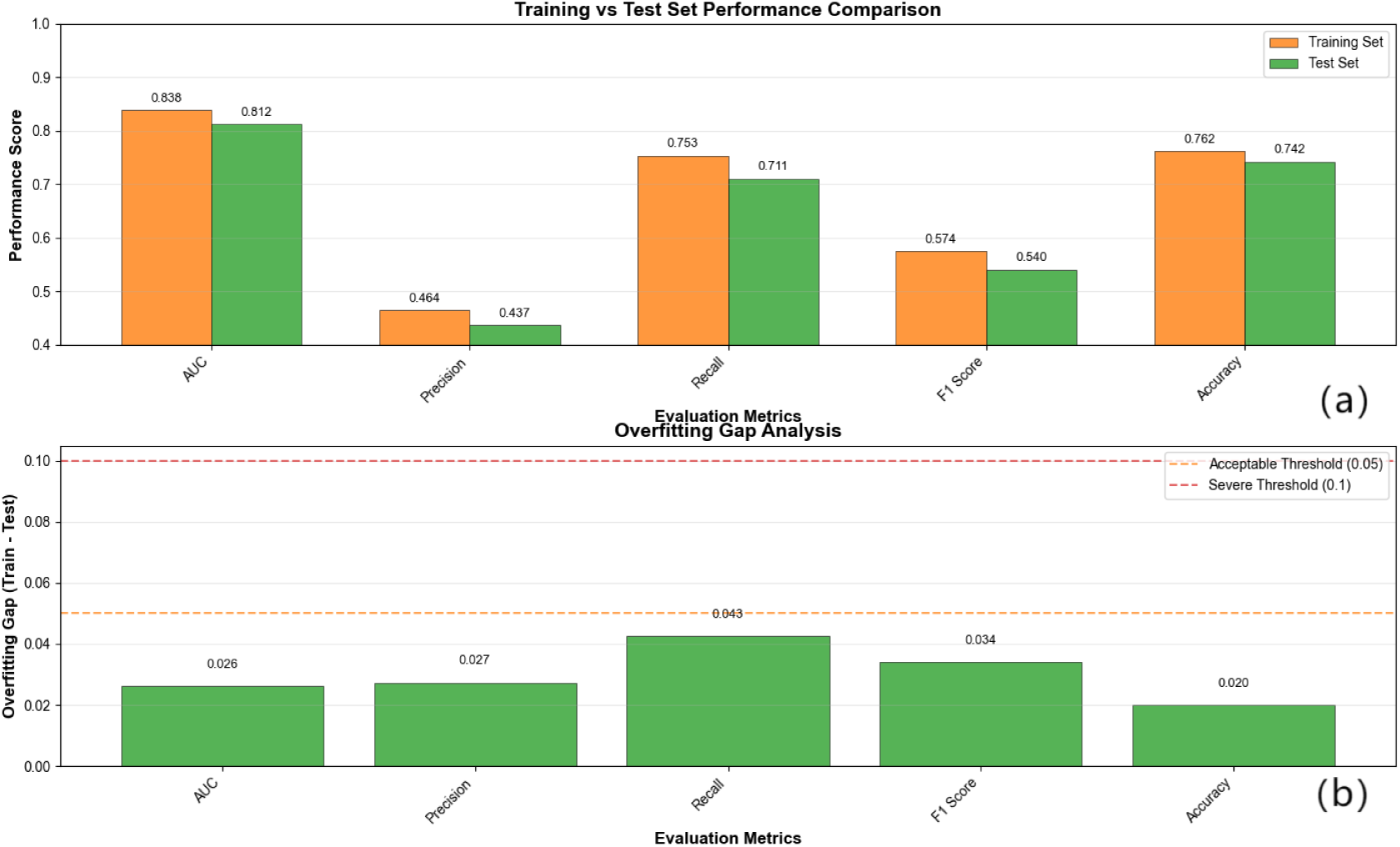
(a) Performance comparison between training and test sets for Model 1. (b) Overfitting detection plot for the training set, illustrating the generalization gap.

**Figure 5.**
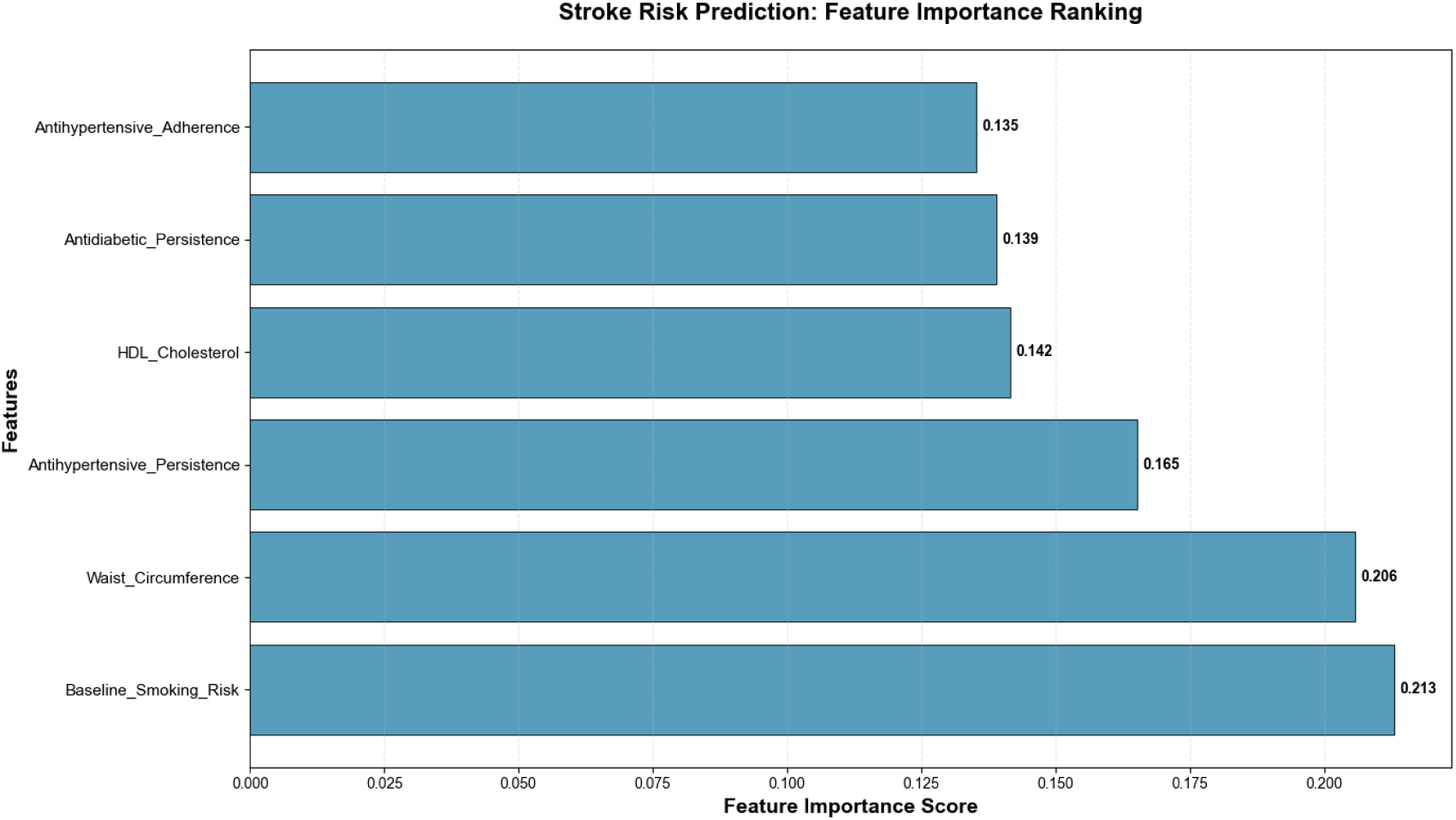
Bar plot of core feature importance in Model 1 (Random Forest-based prediction of high-risk conversion in the general population).

#### 2.2 Model 2 (Prediction of Stroke Onset in High-Risk Populations)

Among the seven candidate algorithms, the conservative-parameter LightGBM (LGB_Conservative) demonstrated the optimal performance:

- It achieved an AUC of 0.8479 in the test set, which was statistically significantly better than the other six algorithms (2 regularized logistic regressions, 2 parametric gradient Random Forests, GBM, and XGBoost).
- The model utilized 12 key features, with the top 5 being: age at medical record creation, homocysteine, number of carotid plaques, fasting blood glucose, and blood glucose change slope. These features enabled precise stratification of short-term stroke onset risk in the high-risk population.
- The overfitting gap (defined as the AUC difference between the training and test sets) was −0.0318, reflecting excellent stability.
- Core hyperparameters of the LGB_Conservative model were: n_estimators = 100, num_leaves = 31, learning_rate = 0.05, max_depth = 5, subsample = 0.7, colsample_bytree = 0.7, reg_alpha = 0.5, reg_lambda = 1.0, class_weight = “balanced”, and min_child_samples = 20.

**Figure 6.**
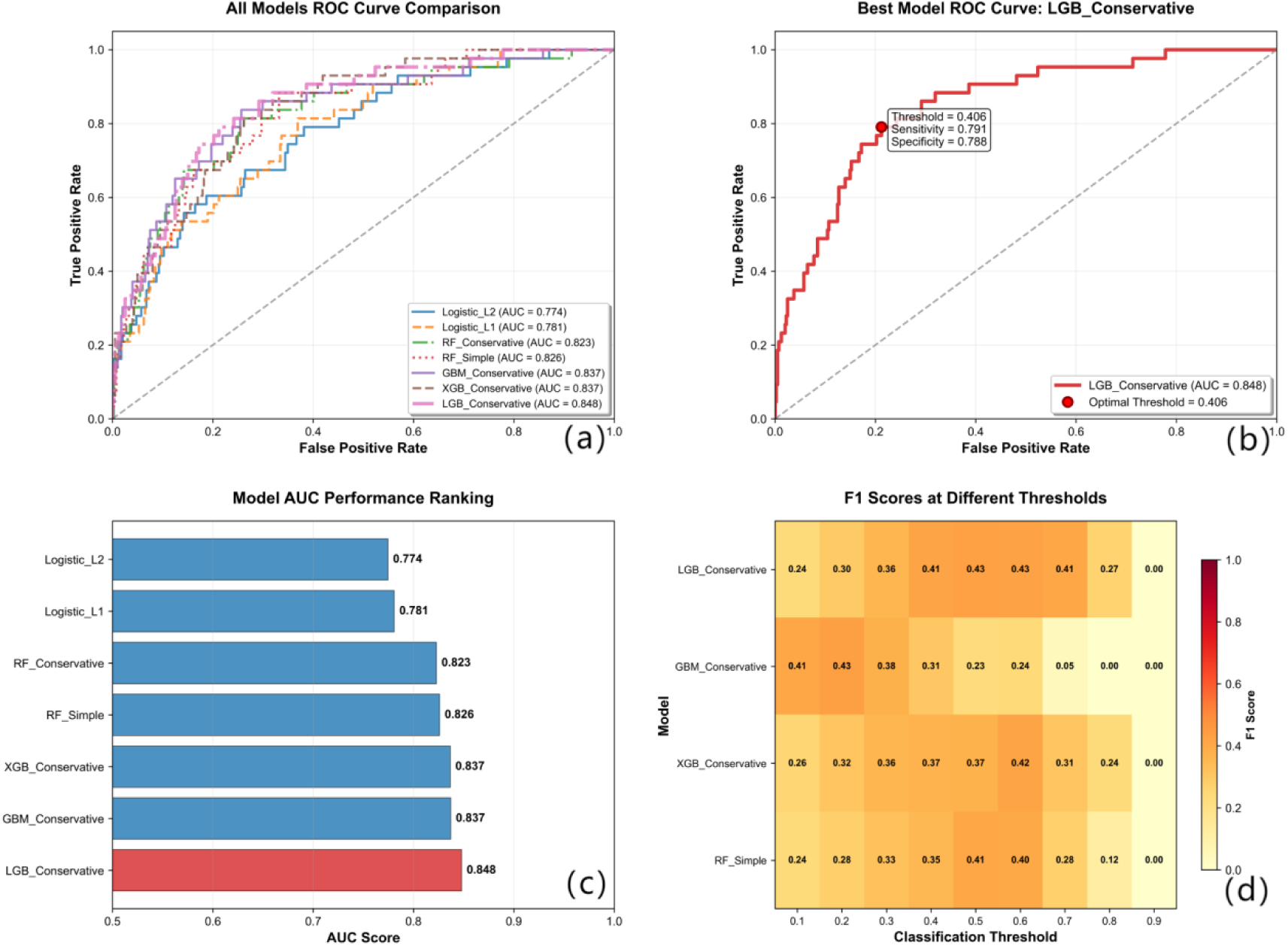
(a) Receiver operating characteristic (ROC) curves of seven candidate algorithms for Model 2 (stroke onset prediction in high-risk populations). (b) Detailed ROC analysis of the optimal model (LGB_Conservative). (c) Bar chart ranking the AUC performance of each model. (d) F1-scores of each model under different classification thresholds.

**Figure 7.**
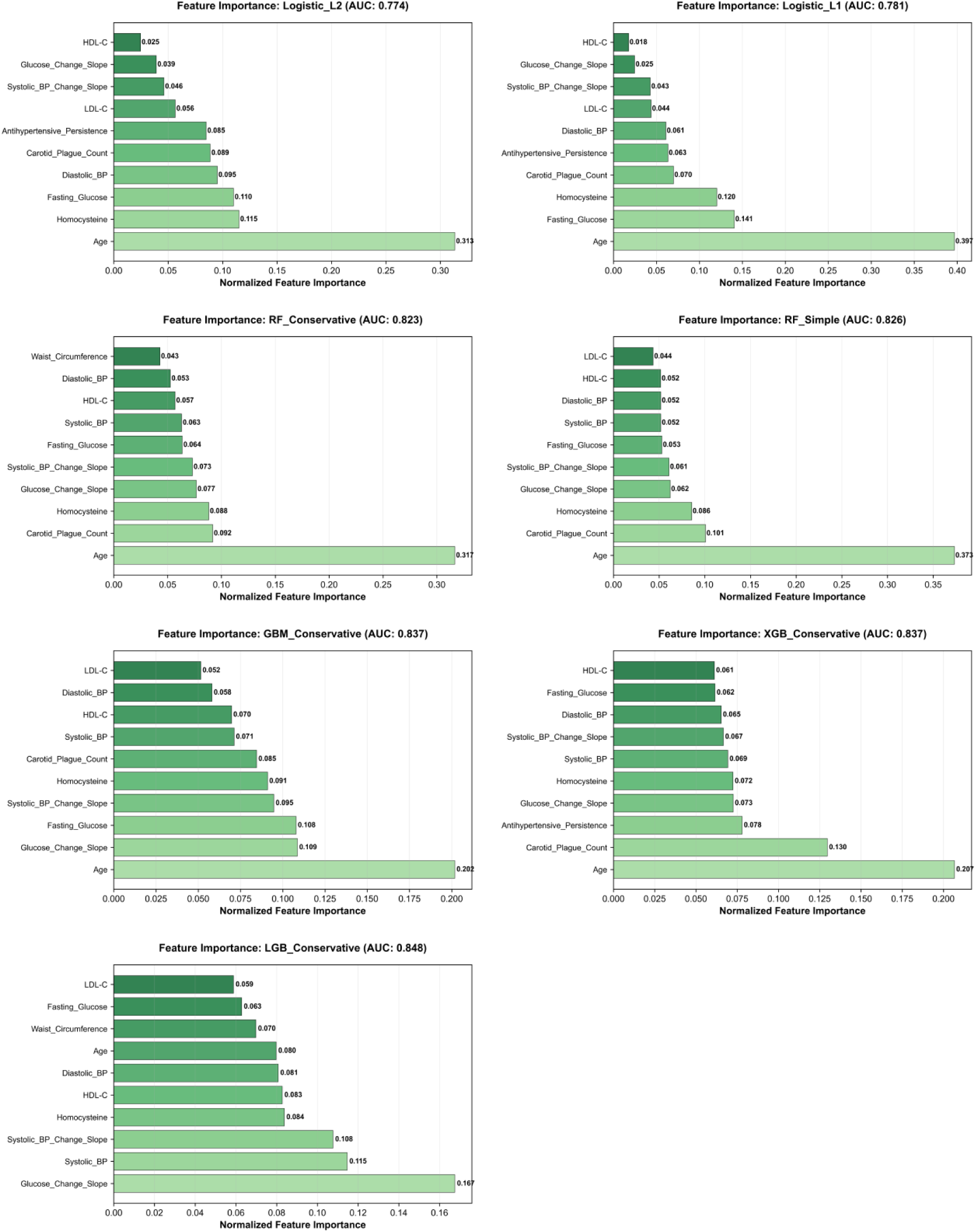
Comparison of feature importance across the seven candidate algorithms in Model 2.

**Figure 8.**
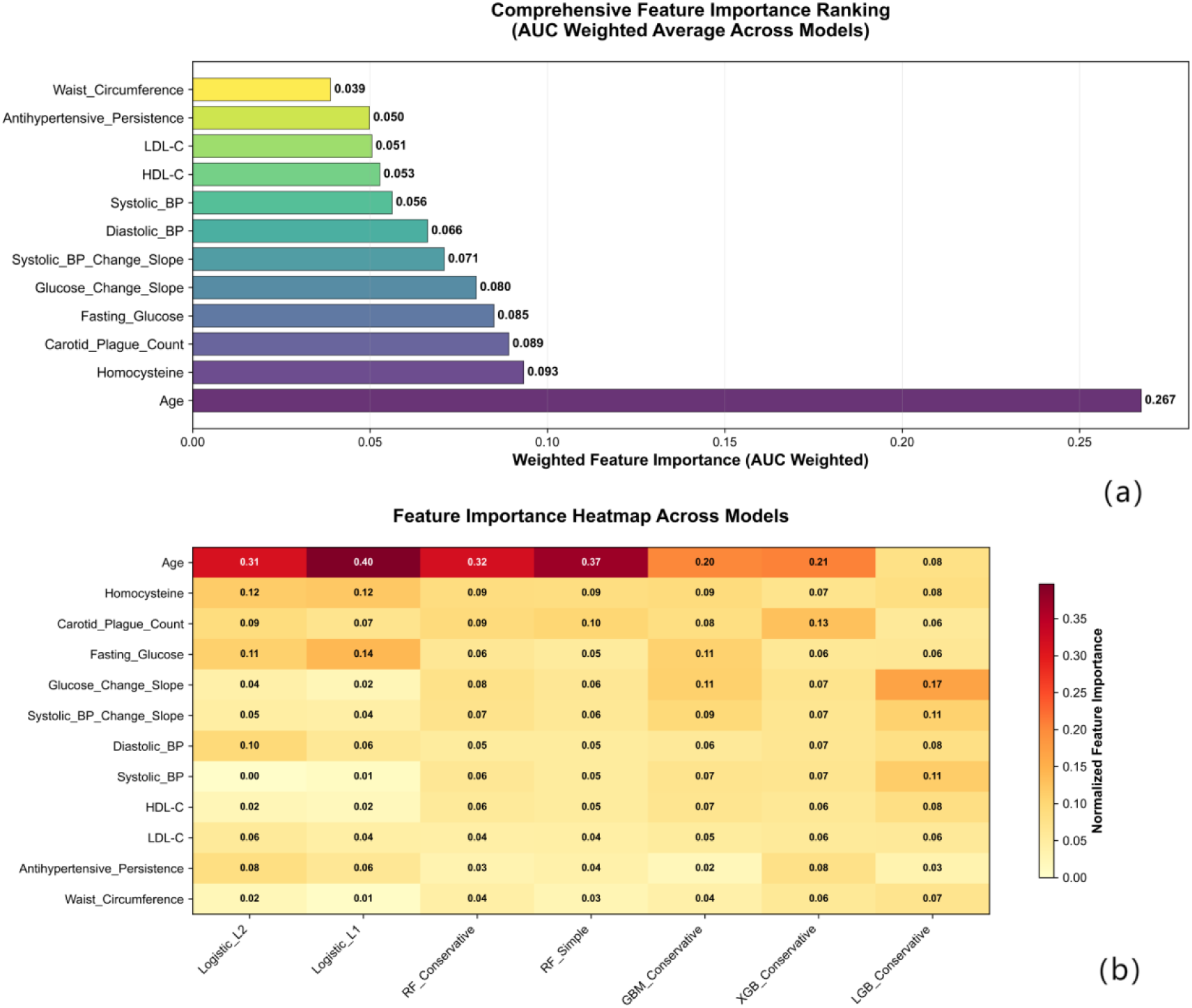
(a) Comprehensive feature importance bar chart for the optimal Model 2 (LGB_Conservative). (b) Heatmap of feature importance across all seven models in Model 2.

##### 2.3 Model 3 (Individual Variability and Trajectory Feature-Oriented Prediction)

Constructed based on 48 trajectory features (including trend slopes, variability indicators, last measurement values, and composite abnormality scores), this model successfully identified 15 core predictors—such as systolic blood pressure variability and last-measured BMI.

- For dual-outcome prediction (high-risk conversion and stroke onset) in the test set, the model achieved an AUC of 0.761 and a Precision-Recall Area Under the Curve (PR-AUC) of 0.489. The PR-AUC was additionally reported to account for the imbalance in outcome events.
- Through risk stratification, the population was categorized into four risk levels: the event rate was only 3.1% in the low-risk group, while it reached 95.2% in the very high-risk group—a difference that was highly statistically significant (P < 0.001).
- Furthermore, the model identified three distinct patterns of abnormal trajectories in 247 high-risk patients: rapid blood pressure elevation type, hypertensive fluctuation type, and composite abnormality type.
- Core hyperparameters of the model were: n_estimators = 200, max_depth = 15, min_samples_split = 20, min_samples_leaf = 10, bootstrap = True, and random_state = 42.

- Importantly, variability indicators (e.g., systolic blood pressure variability) were confirmed as independent predictive factors, which enhanced the ability to identify potentially high-risk individuals with “normal absolute values but abnormal trajectories”—a population often missed by static models.

**Figure 9.**
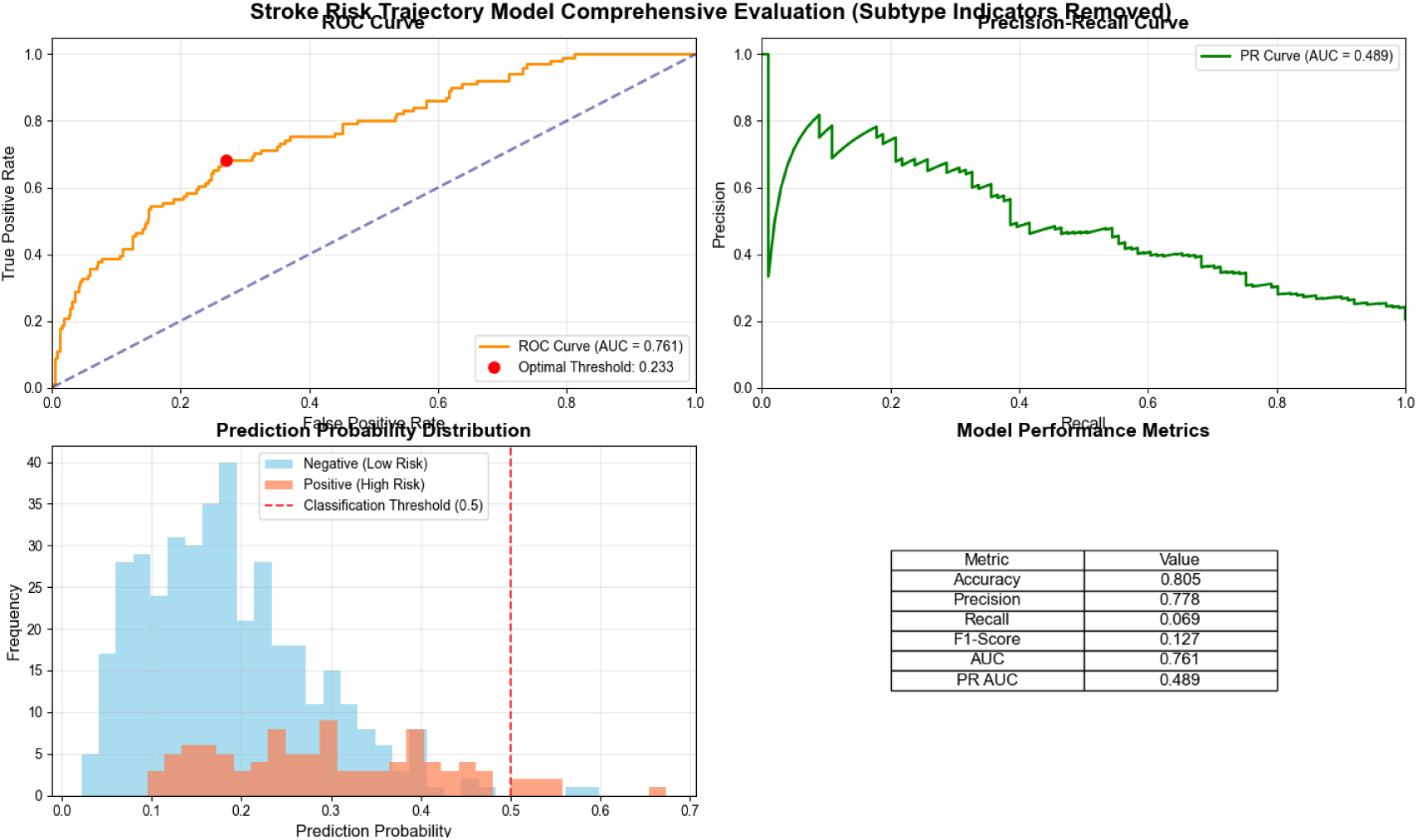
Comprehensive evaluation of Model 3 (trajectory-based prediction for longitudinal populations), including ROC curve, precision-recall (PR) curve, and predicted probability distribution.

**Figure 10.**
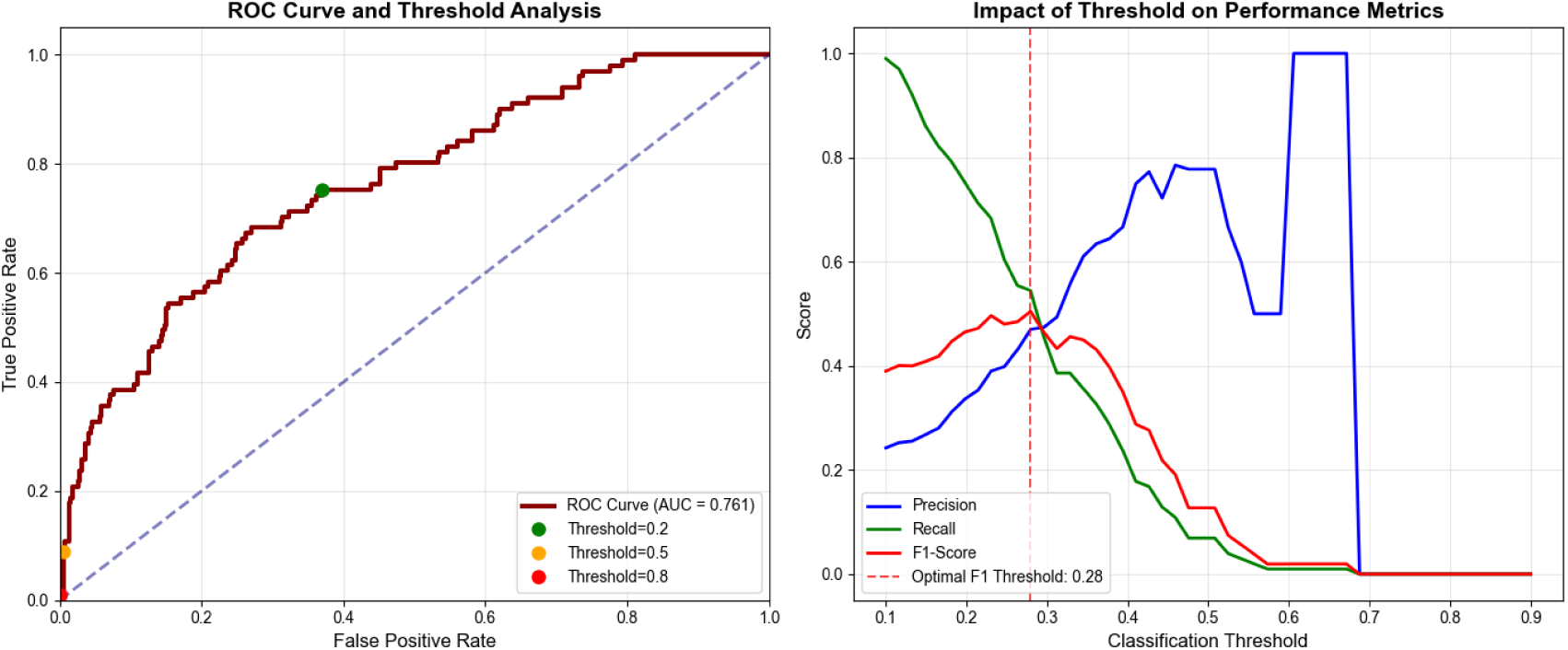
Detailed ROC analysis for Model 3, demonstrating the impact of varying classification thresholds on performance metrics.

**Figure 11.**
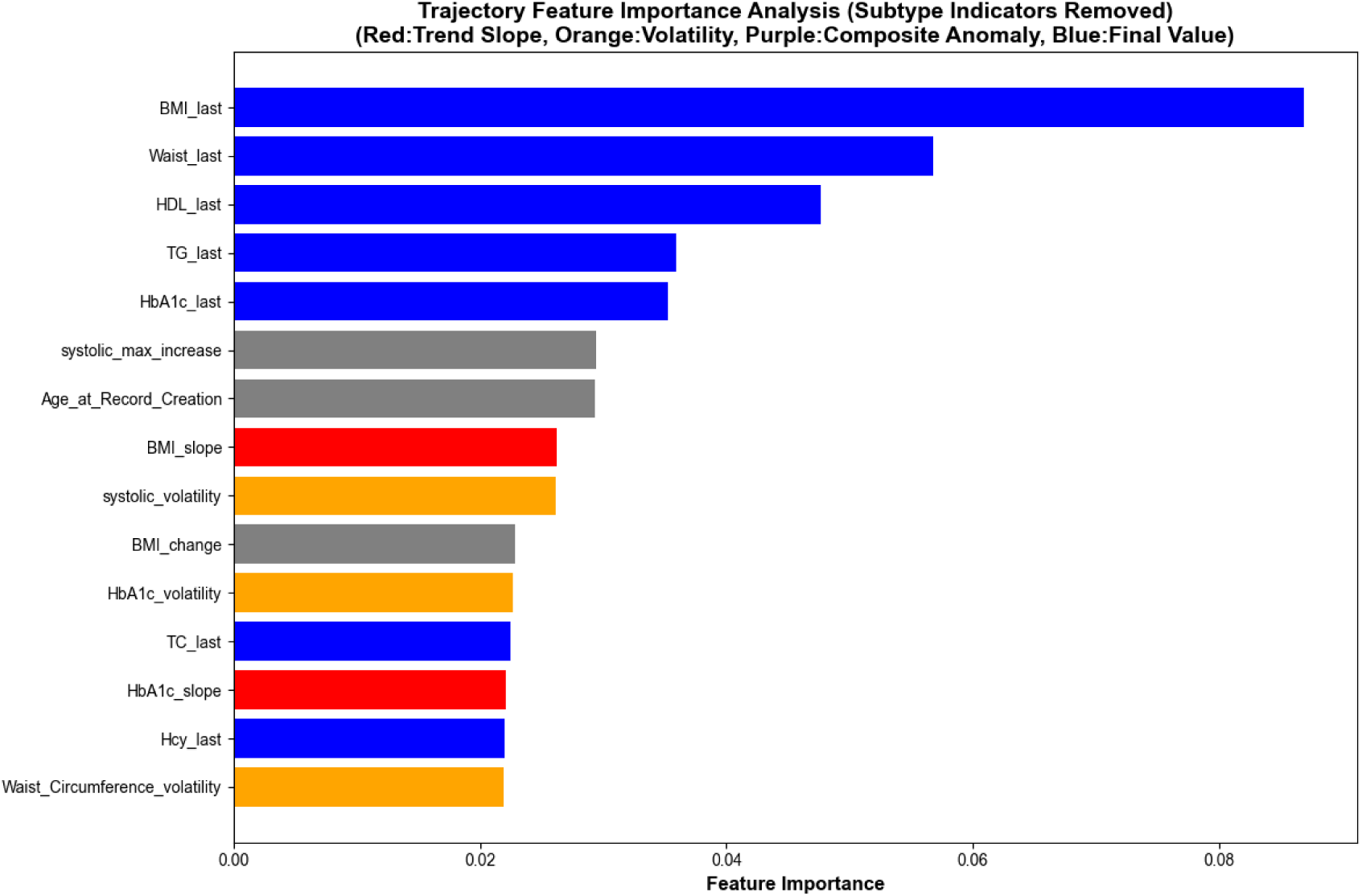
Bar chart of feature importance in Model 3 (Random Forest model incorporating 48 trajectory features).

## Discussion

This study constructed and validated three stratified machine learning models for initial screening of the general population, precision screening of high-risk groups, and monitoring of longitudinal populations based on data from a single-center longitudinal prospective cohort, forming a logically progressive system for stroke risk prediction. The core breakthrough of this system lies in overcoming the inherent limitation of traditional static models that rely on cross-sectional data—by integrating dynamic physiological parameters and behavioral indicators, it achieves precise risk stratification for populations at different risk stages, providing a clinically applicable tool for primary stroke prevention.

From the perspective of optimal algorithm adaptability:

- For initial screening of the general population, the Random Forest algorithm was selected as the optimal model, as it can naturally fit the non-linear relationships between behavioral variables (e.g., Basic Smoking Risk Score) and metabolic variables (e.g., waist circumference) and exhibits strong robustness to multi-type features (behavioral, static, dynamic). This model ultimately retained 6 core features, enabling large-scale screening in primary medical institutions without complex data preprocessing. It not only balances “simplicity” and “accuracy” but also significantly reduces the missed detection of potentially high-risk individuals with “normal static indicators but abnormal dynamic trends,” which aligns with the practical needs of community-based initial screening.
- For precision screening of high-risk populations, LGB_Conservative (LightGBM with conservative parameters) was identified as the optimal algorithm. Addressing the small-sample imbalance issue (stroke event rate of only 9.8%), this algorithm effectively controlled overfitting through dual regularization (reg_alpha = 0.5, reg_lambda = 1.0) and sample/feature sampling strategies (subsample = 0.7, colsample_bytree = 0.7). Among the 12 key features it integrated, the top 5 (age at medical record creation, homocysteine, number of carotid plaques, fasting blood glucose, slope of blood glucose change) can accurately distinguish between short-term and long-term onset risks within the high-risk population, resolving the problem that traditional static models fail to identify risk heterogeneity in high-risk groups and providing a basis for the optimal allocation of preventive resources.
- For longitudinal population monitoring, a trajectory feature-oriented Random Forest model was adopted. The 48 trajectory features (including trend slopes, variability indicators, last measurement values, and composite abnormality scores) can simultaneously capture long-term metabolic homeostasis and abnormal parameter fluctuations, meeting the dual goals of “risk early warning” and “intervention effect evaluation.” It serves as an irreplaceable tool for dynamic monitoring in chronic disease management.

The ranking of core feature importance is highly consistent with the risk-driven mechanisms of different populations, collectively constructing a “feature-target” system for stratified stroke prevention:

- The initial screening model for the general population focuses on the “behavior-metabolism” dual core: The Basic Smoking Risk Score quantifies cumulative tobacco exposure and risk attenuation after smoking cessation, reflecting long-term tobacco-related risk more accurately than the traditional “smoker/non-smoker” binary classification; waist circumference is more effective than BMI in identifying “potentially high-risk individuals with normal BMI but excessive visceral fat,” which aligns with the clinical recognition of abdominal obesity as a core marker of metabolic syndrome^[18]^; the persistence of antihypertensive/hypoglycemic treatment and antihypertensive adherence grades extend risk assessment from the “disease diagnosis level” to the “treatment management level,” filling the gap of traditional models that “prioritize diagnosis over management”; high-density lipoprotein cholesterol (HDL-C), as a protective indicator^[20]^, complements the dimension of metabolic risk assessment.
- The precision screening model for high-risk populations uses age at medical record creation as the basic risk anchor. With increasing age, vascular degenerative changes combined with underlying diseases lead to an exponential increase in stroke risk, which is consistent with the age-related risk patterns reported in the Global Burden of Disease Study^[21]^; homocysteine, as a subclinical risk marker, can identify individuals with “no obvious symptoms but impaired vascular endothelium,” and its elevated levels exhibit synergistic risk with hypertension and diabetes^[22]^; the number of carotid plaques directly indicates atherosclerotic burden, providing a pathological basis for clinically deciding whether to initiate intensive lipid-lowering and antiplatelet therapy**Error! Reference source not found.**; the combination of fasting blood glucose and slope of blood glucose change covers both static metabolic levels and dynamic trend information, avoiding misjudgment based solely on a single blood glucose measurement ^[23]^.
- The longitudinal population monitoring model uses last-measured metabolic indicators (e.g., last BMI, last waist circumference, last HDL-C) as the quantitative basis for long-term intervention effects—for example, if an individual’s waist circumference decreases from an excessive level to the normal range during follow-up, the last measurement value can directly reflect the degree of risk reduction; the inclusion of indicators such as systolic blood pressure variability (ranked 9th in importance)^[25]^ and glycated hemoglobin variability (ranked 11th in importance) upgrades risk assessment from “indicator value compliance” to “indicator fluctuation stability,” effectively avoiding the missed detection of individuals with “normal values but abnormal trajectories” by traditional models.

The core innovation of this study lies in the systematic integration of variability and dynamic trend indicators into the prediction framework:

- The long-term coefficient of variation of systolic blood pressure in Model 1 can identify “latently high-risk individuals with normal single-measurement blood pressure but large long-term fluctuations.” Such fluctuations increase stroke risk by activating vascular endothelial injury pathways and accelerating atherosclerosis^[26]^. Primary medical institutions can calculate this indicator using only 3 follow-up blood pressure measurements without additional testing costs, significantly improving the accuracy of community-based screening.
- The slope of blood glucose change and slope of systolic blood pressure change in Model 2 establish a “dynamic trend evaluation dimension” for high-risk populations: Individuals with fasting blood glucose at the upper normal limit (5.6–6.1 mmol/L) but a continuously rising slope have a much higher stroke risk than those with stable indicators (Gerbaud et al., 2019) [9]. Moreover, the synergistic effect between these slope indicators and the number of carotid plaques can reflect the “effect of metabolic fluctuations accelerating pathological progression,” providing a clear pathway for formulating clinical intervention strategies of “first stabilizing indicator fluctuations, then controlling plaque progression.”
- The systolic blood pressure variability ^[27]^ and glycated hemoglobin variability indicators in Model 3 shift the evaluation of intervention effects in longitudinal populations from “static compliance” to “dynamic stability”—some patients may meet clinical standards for indicator values but still have higher-than-normal variability, and their stroke risk remains significantly higher than that of individuals with normal fluctuations^[26]^. Such risks can only be identified through variability indicators, which also upgrades this model from a simple “risk prediction tool” to an “intervention effect monitoring tool.”

Compared with traditional static models, the improved predictive performance of this system relies on multi-variable input (e.g., 12 features in Model 2, 48 trajectory features in Model 3). However, with the advancement of electronic health record (EHR) systems, clinical workflows have realized the automatic extraction of longitudinal data (e.g., multiple blood pressure measurements, medication adherence records) and real-time feature calculation, which not only reduces the workload of manual variable entry but also lowers the reliance on model “simplicity.” In addition, machine learning models (such as Random Forest and LGB_Conservative used in this study) can be retrained and optimized using additional multi-center data, showing promising prospects for enhancing the generalizability of the models to populations in different regions (e.g., rural vs. urban) and ethnic groups.

This study has several limitations: (1) The single-center design—data were derived from the longitudinal prospective cohort in Qingshanhu District, Nanchang City—so external validation using multi-center and multi-ethnic cohort data is required to confirm the generalizability of the models; (2) Differences in variable availability may affect model application—for example, some primary medical institutions may lack the carotid ultrasound data required for Model 2 or the complete longitudinal trajectory data required for Model 3, so adjustments based on variable availability are needed when integrating multi-center data; (3) The study excluded patients with incomplete follow-up (fewer than 3 follow-up visits) and those who received endovascular recanalization therapy: the former exclusion was to ensure the reliability of trajectory features, while the latter was because treatment-related variables (e.g., recanalization success rate) are only applicable to this subgroup, resulting in the models being temporarily unsuitable for populations undergoing acute interventional therapy; (4) The interactive effects of key features (e.g., synergistic risk between Basic Smoking Risk Score and waist circumference) were not explored in depth; future studies should further investigate risk mechanisms using interpretable artificial intelligence technologies.

### Conclusion

This study demonstrates that the multi-tiered dynamic stroke risk prediction system constructed based on data from a single-center longitudinal prospective cohort—comprising three logically progressive machine learning models (Random Forest for initial screening of the general population, LGB_Conservative for precision screening of high-risk populations, and trajectory feature-oriented model for longitudinal populations)—effectively compensates for the limitations of traditional static models (which rely on cross-sectional data and fail to capture dynamic risks) and achieves precise risk stratification for different populations.

The core value of this system lies in integrating the variability of dynamic physiological parameters (e.g., systolic blood pressure variability) and behavioral indicators (e.g., treatment adherence), which significantly improves the ability to identify potentially high-risk individuals with “normal static indicators but abnormal trajectories” and provides clinically applicable tools for primary screening, high-risk precision screening, and longitudinal monitoring.

It should be noted that this study is a single-center research, so the generalizability of the models needs to be verified using multi-center data. Nevertheless, this multi-tiered dynamic prediction system provides a scientific and feasible new paradigm for personalized stroke prevention and is expected to promote the transformation of primary stroke prevention toward precision and individualization.

## Author Contributions

Conceptualization: Zhirui Liao;

Data Curation: Jing Huang, Ximei Chen, Yinghua Yang;

Formal Analysis: Zhirui Liao;

Funding Acquisition:Yinghua Yang;

Investigation: Jing Huang, Ximei Chen;

Methodology: Zhirui Liao;

Project Administration: Zhirui Liao, Yinghua Yang;

Resources: Jing Huang, Ximei Chen, Yinghua Yang;

Software: Zhirui Liao;

Validation: Zhirui Liao, Zesheng Yan;

Visualization: Zhirui Liao, Zesheng Yan;

Writing - Original Draft: Zhirui Liao;

Writing - Review & Editing: All authors.

## Data Availability

The datasets supporting the findings of this study are derived from a single-center longitudinal prospective cohort (2018-2022) of community-dwelling populations and outpatients enrolled by the Center for Disease Control and Prevention of Qingshanhu District, Nanchang City, and the School of Public Health, Wuhan University. These datasets contain potentially sensitive personal health information and are subject to the ethical approval requirements of the Ethics Committee of Xuanwu Hospital, Capital Medical University (Approval No.: Clinical Research Review [2015] No. 024). Therefore, the datasets are not publicly available to protect participant confidentiality. Qualified researchers may request access to the de-identified data by contacting the corresponding authors (Yinghua Yang; Zhirui Liao) with a detailed research proposal. Data access will be granted following review to ensure compliance with ethical guidelines and participant privacy protection. No external datasets or supplementary materials hosted in public online repositories were used in this study, so no relevant URLs are provided.

## Acknowledgements

The authors would like to express sincere gratitude to Prof. JUNYONG ZHU (School of Public Health, Wuhan University) for his valuable contributions to this study, including the design and direction of the overall research framework, professional guidance on the technical route optimization, and critical revision of the manuscript content.

